# Perceptions of a Menthol Cigarette and Flavored Cigar Ban in Black and White Adults in the United States who Smoke Menthol Cigarettes and Factors Associated with Ban Opposition or Ambivalence

**DOI:** 10.1101/2025.04.18.25326088

**Authors:** Michael J. Arnold, Eleanor L.S. Leavens, Lisa Sanderson Cox, Alexandra Brown, Matthew S. Mayo, Nathaniel L. Baldwin, Thu A. Nguyen, Nicole L. Nollen

**Affiliations:** Department of Population Health, University of Kansas School of Medicine, Kansas City, KS; University of Kansas Cancer Center, University of Kansas School of Medicine, Kansas City, KS; Department of Biostatistics and Data Science, University of Kansas School of Medicine, Kansas City, KS

**Keywords:** Black or African American, Health Disparities, Health Policy, Social Determinants of Health, Tobacco

## Abstract

**Objectives:** Menthol flavoring is a critical public health issue, but prior work has largely represented the voices of White adults who smoke (AWS) menthol cigarettes who comprise a small subset of AWS menthol cigarettes in the US. This study compared perceptions of a hypothetical MC/FC ban among Black and White AWS menthol cigarettes.

**Methods:** Participants were a convenience sample of 2,113 Black and 1,087 White AWS menthol cigarettes collected through Amazon Mechanical Turk between July 2023 and January 2024. Participants reported opinions about a MC/FC ban, likely public health outcomes, and hypothetical impact of the ban on their smoking behavior. Stepwise logistic regression modeled factors associated with ban opposition/ambivalence.

**Results:** Over one-third of menthol cigarette users supported a MC/FC ban (Black, 37.2% vs White, 34.5%, p=.13], but Black AWS were more likely to endorse public health benefits of a ban assessed via agreement with 5 statements of FDA rationale [3.0 (SD=1.7) versus 2.4 (SD=1.8), p<.001]. Smoking more cigarettes per day, belief that menthol cigarettes are more addictive/harder to quit, and intent to continue using nicotine under a ban increased odd of opposition/ambivalence.

**Conclusions:** Compared to White AWS, Black AWS were more likely to believe that a MC/FC ban would benefit public health and showed no statistical difference in overall support for a ban. Targeted outreach to those who consume more menthol products and those who do not intend to quit nicotine could increase ban support among menthol users.

**Summary Box:** *What is the current understanding of this subject?:* Knowledge of the public’s perception of a MC/FC ban is largely informed by White menthol cigarette users. Given that Blacks represent the majority of AWS menthol cigarettes in the US, the paucity of data from Black voices engenders a critical gap in research used to inform health policy.

*What does this report add to the literature?:* Compared to White AWS, Black AWS were more likely to believe that a MC/FC ban would benefit public health and showed no statistical difference in overall support for a ban. To our knowledge, this study comprises the largest convenience sample of perceptions and perceived impact of a federal MC/FC product standard among Black AWS menthol cigarettes in the US and provides valuable evidence to inform policy action in these areas.

*What are the implications for public health practice?:* Findings support ongoing efforts to advance a MC/FC product standard and suggest that targeted outreach to those who consume more menthol products and those who do not intend to quit nicotine could increase ban support among menthol users.

## INTRODUCTION

Menthol as a characterizing flavor in cigarettes is a critical public health issue due, in large part, to targeted tobacco industry marketing that has led to a high number of Black adults using menthol cigarettes, which are more addictive and harder to quit.^1-4^ Currently, 81% of Black adults who smoke (AWS) and 34%-51% of AWS in other racial and ethnic minority groups use menthol cigarettes.^5^ Further, the overall number of adults who use menthol cigarettes has been increasing since 2008, reaching 43% in 2020.^5-6^

The US Food and Drug Administration (FDA) gained authority to regulate tobacco products in 2009 under the Family Smoking Prevention and Tobacco Control Act.^7^ This law eliminated all flavored cigarettes, except menthol, and formed the Tobacco Products Scientific Advisory Committee (TPSAC), which was charged with investigating the threat of menthol cigarettes to public health. Following extensive review, TPSAC concluded that “removal of menthol cigarettes from the marketplace would benefit public health in the United States.”^8^ Simulation modeling estimates that a menthol cigarette ban implemented in 2021 would save over a quarter million Black lives by 2060.^9^ To-date, no federal menthol cigarette or flavored cigar (MC/FC) product standards (commonly referred to as ‘bans’) have been enacted, despite continued evidence that these actions have the potential to improve public health by reducing cigarettes per day (CPD) and promoting smoking cessation among AWS menthol cigarettes.^10-12^

Recent prominent news stories cite fear of backlash from the Black community as one reason for failure to enact a MC/FC product standard^13^ despite overwhelming public support for a MC/FC product standard from Black leaders and other health care groups.^12,14-16^ However, the opinions of Black AWS menthol cigarettes are substantially underrepresented in current research. While existing studies provides valuable information about perceptions of a MC/FC ban among menthol cigarette users ^16-25^ and its impact on their tobacco use behaviors,^11,18-19,24-25^ findings overwhelmingly represent those of White AWS menthol cigarettes. Given that Blacks represent the majority of AWS menthol cigarettes in the US, the paucity of data from Black voices engenders a critical gap in research used to inform health policy.^5^

To address this gap, the current study 1) compared perceptions of a MC/FC ban among Black and White AWS menthol cigarettes and 2) examined demographic, tobacco use, and other factors associated with opposition or ambivalence to the ban. A menthol cigarette and flavored cigar product standard was assessed together because they will both disproportionately impact Black adults who are more likely to consume these products. While a MC/FC product standard was withdrawn from the Office of Management and Budget register on January 2025, it was active during the time the study was conducted and remains highly relevant as momentum for implementing MC/FC bans at the state and local level continues to increase.^26^ To our knowledge, this study comprises the largest convenience sample of perceptions and perceived impact of a MC/FC product standard among Black AWS menthol cigarettes in the US and provides valuable evidence to inform policy action in these areas.

## METHODS

Participants were registered workers on Amazon’s Mechanical Turk (MTurk), an online crowdsourcing platform that has been used widely in tobacco control research, who self-opted into a study titled “Smoking & Health Behaviors Survey” between July 2023 and January 2024. Interested respondents completed informed consent and a screening questionnaire. Eligible participants were African American/Black or White adults (≥ 21 years of age) who had been menthol cigarette smokers for at least one year and were using a computer with a United States IP address. Participants were excluded if they were primary users of non-cigarette tobacco products or lived in a place where the sale of menthol cigarettes was prohibited (e.g., California, Massachusetts). Using standard methods, those who incorrectly answered ≥ 2 of 3 validity questions placed throughout the survey or who indicated ‘do not use my data’ were excluded.^27^

The study was administered anonymously through REDCap and approved by the University of Kansas Medical Center’s Institutional Review Board (STUDY00160222), and its conduct was consistent with applicable federal law and CDC policy. Participants were compensated $5 for completion of the survey. Surveys took about 10 minutes to complete.

The flow of participants into the study is shown in Figure 1; 6,924 potential participants were screened to enroll the targeted sample size of 3,200 participants (2,133 Black, 1087 White).

### Measures

Exact survey questions and response options are outlined in Tables 1-4. Questions and response options were adapted by the research team from the European Union’s (EU) 2020-2021 International Tobacco Control Survey.^28^

**Table 1.**
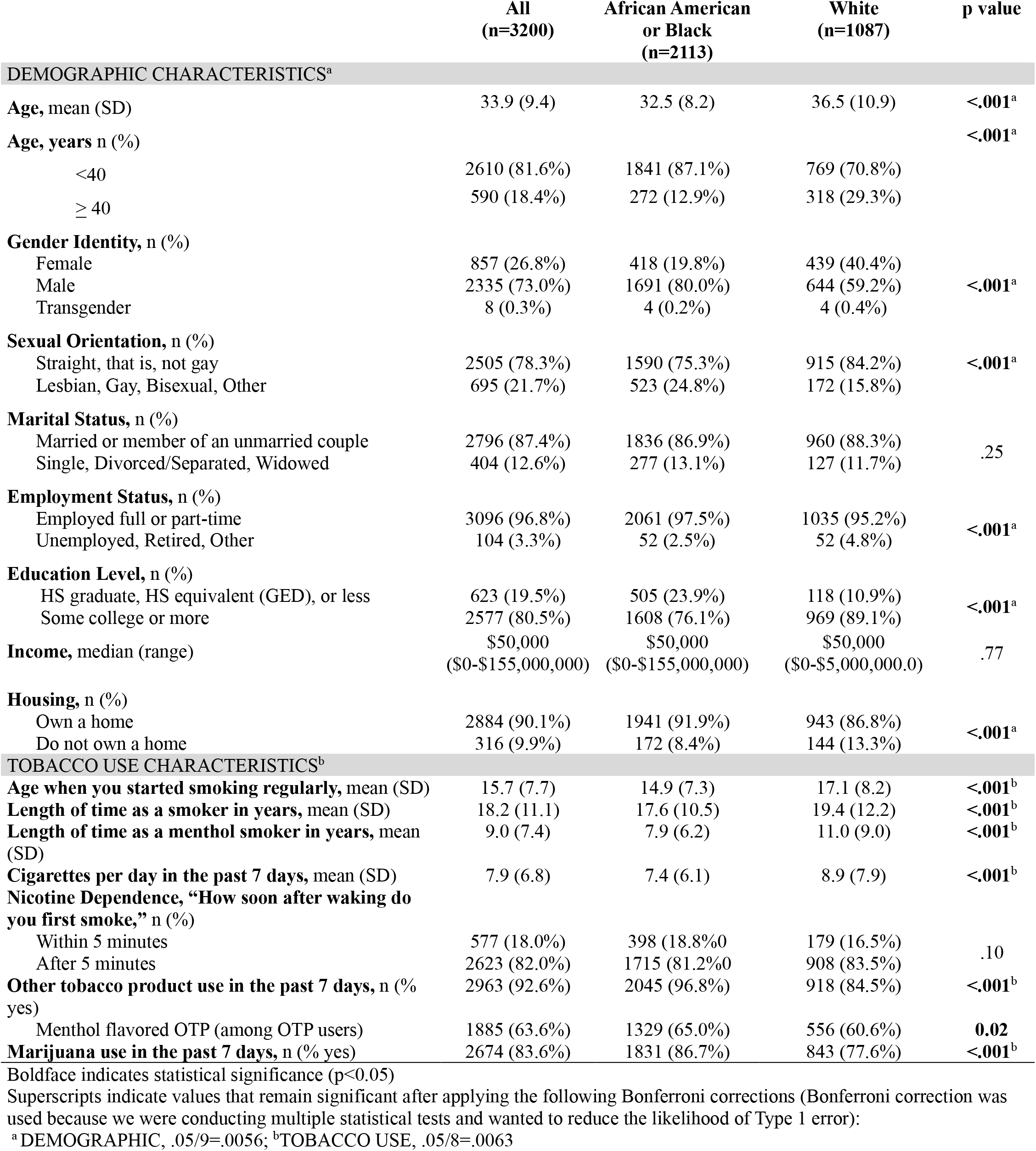
Comparison of Demographic and Tobacco Use Characteristics by Race among AWS Menthol Cigarettes.

#### Demographic and tobacco use characteristics

Participants reported their age, gender identity, race, sexual orientation, marital status, employment status, education level, income, home ownership, length of time as a smoker overall and as a menthol smoker, age started smoking regularly, cigarettes per day (CPD), nicotine dependence (FTND), and use of other tobacco products (OTP) and marijuana (MJ).

#### Beliefs about menthol cigarettes versus non-menthol cigarettes

Three questions assessed participants beliefs about the addictiveness, harmfulness, and difficulty in quitting menthol cigarettes versus non-menthol cigarettes.

#### Opinions and perceived public health outcomes

A brief statement outlined the FDA’s proposed product standard and explained that menthol cigarettes and flavored cigars would not be available for purchase if the product standards were enacted.^29^ A single item assessed participants’ knowledge of the MC/FC ban before completing the survey. Follow-up items gauged participants’ personal feeling about a federal ban and belief about their community’s feelings about a ban. Those in favor and opposed indicated the primary reason for their support or opposition, respectively (Supplementary Table 1). Five statements outlined the FDA’s suggested public health benefits of a MC/FC ban (Table 2, e.g., a ban ‘would prevent youth from ever starting smoking,’ ‘encourage menthol smokers to smoke less,’). Participants indicated their belief about the likelihood of each outcome as a result of a ban.^29^

**Table 2.** Comparison of Beliefs and Opinions about a MC/FC Ban Opinions by Race among AWS Menthol Cigarettes.

|  | All<br>(n=3200) | African American<br>or Black<br>(n=2113) | White<br>(n=1087) | p value |
| --- | --- | --- | --- | --- |
| **BELIEFS ABOUT MENTHOL CIGARETTES<sup>c</sup>** |  |  |  |  |
| **Addictiveness of menthol vs. non-menthol cigarettes, n (%)** |  |  |  |  |
| Less or equally addictive | 1658 (51.8%) | 974 (46.1%) | 684 (63.0%) | <.001<sup>c</sup> |
| More addictive | 1541 (48.2%) | 1139 (53.9%) | 402 (37.0%) |  |
| **Harmfulness of menthol vs. non-menthol cigarettes, n (%)** |  |  |  |  |
| Less or equally as harmful | 1941 (60.7%) | 1228 (58.1%) | 713 (65.7%) | <.001<sup>c</sup> |
| More harmful | 1258 (39.3%) | 885 (41.9%) | 373 (34.4%) |  |
| **Difficulty to quit of menthol vs. non-menthol cigarettes, n (%)** |  |  |  |  |
| Less or equally as hard to quit | 1928 (60.3%) | 1190 (56.3%) | 738 (68.0%) | <.001<sup>c</sup> |
| Harder to quit | 1271 (39.7%) | 923 (43.7%) | 348 (32.0%) |  |
| **Overall beliefs about Menthol Summary: Out of the 3 statements regarding beliefs about menthol cigarettes, how many thought menthol was more addictive, more harmful, harder to quit, mean (SD)** | 1.3 (1.1) | 1.4 (1.1) | 1.0 (1.0) | <.001<sup>c</sup> |
| **OPINIONS OF THE PROPOSED MENTHOL AND FLAVORED CIGAR PRODUCT STANDARD<sup>d</sup>** |  |  |  |  |
| **Before today, had you heard about a possible ban on menthol cigarettes and all flavored cigars?, n (%)** |  |  |  |  |
| Yes | 2799 (87.5%) | 1933 (91.5%) | 866 (79.7%) | <.001<sup>d</sup> |
| No | 401 (12.5%) | 180 (8.5%) | 221 (20.3%) |  |
| **What are your feelings about a ban on menthol cigarettes and flavored cigars?, n (%)** |  |  |  |  |
| Opposed | 1122 (35.1%) | 716 (33.9%) | 406 (37.4%) | 0.13 |
| Ambivalent | 917 (28.7%) | 611 (28.9%) | 306 (28.2%) |  |
| In favor | 1161 (36.3%) | 786 (37.2%) | 375 (34.5%) |  |
| **In your opinion, how would others in your community feel about a ban on menthol cigarettes and flavored cigars?, n (%)** |  |  |  |  |
| Opposed or ambivalent | 1883 (58.8%) | 1274 (60.3%) | 609 (56.0%) | .02 |
| In favor | 1317 (41.2%) | 839 (39.7%) | 478 (44.0%) |  |
| **How likely do you think each of the following things is to happen if the FDA bans menthol cigarettes and flavored cigars?, n (%)** |  |  |  |  |
| **Prevent youth from ever starting smoking** |  |  |  |  |
| Unlikely outcome of the FDA ban | 1101 (34.4%) | 582 (27.5%) | 519 (47.8%) | <.001<sup>d</sup> |
| Likely outcome of the FDA ban | 2099 (65.6%) | 1531 (72.5%) | 568 (52.3%) |  |
| **Encourage menthol smokers to smoke less** |  |  |  |  |
| Unlikely outcome of the FDA ban | 1336 (41.8%) | 810 (38.3%) | 526 (48.4%) | <.001<sup>d</sup> |
| Likely outcome of the FDA ban | 1864 (58.3%) | 1303 (61.7%) | 561 (51.6%) |  |
| **Encourage menthol smokers to quit** |  |  |  |  |
| Unlikely outcome of the FDA ban | 1487 (46.5%) | 913 (43.2%) | 574 (52.8%) | <.001<sup>d</sup> |
| Likely outcome of the FDA ban | 1713 (53.5%) | 1200 (56.8%) | 513 (47.2%) |  |
| **Lead to fewer smoking-related deaths and improve public health** |  |  |  |  |
| Unlikely outcome of the FDA ban | 1500 (46.9%) | 935 (44.3%) | 565 (52.0%) | <.001<sup>d</sup> |
| Likely outcome of the FDA ban | 1178 (55.8%) | 1178 (55.8%) | 522 (48.0%) |  |
| **Improve the health of communities most impacted by menthol smoking, including Black, LGBTQ+, people with mental health conditions, low income/education groups, etc.)** |  |  |  |  |
| Unlikely outcome of the FDA ban | 1531 (47.8%) | 921 (43.6%) | 610 (56.1%) | <.001<sup>d</sup> |
| Likely outcome of the FDA ban | 1669 (52.2%) | 1192 (56.4%) | 477 (43.9%) |  |
| **Overall support for FDA rationale: Out of the 5 statements regarding potential outcomes of a ban on menthol cigarettes and flavored cigars, how many were endorsed as very or somewhat likely, mean (SD)** | 2.8 (1.8) | 3.0 (1.7) | 2.4 (1.8) | <.001<sup>d</sup> |

Boldface indicates statistical significance (p<0.05)
Superscripts indicate values that remain significant after applying the following Bonferroni corrections (Bonferroni correction was used because we were conducting multiple statistical tests and wanted to reduce the likelihood of Type 1 error):
<sup>c</sup>BELIEFS, .05/5=.01; <sup>d</sup>OPINIONS, .05/9=.0056

**Table 3.**
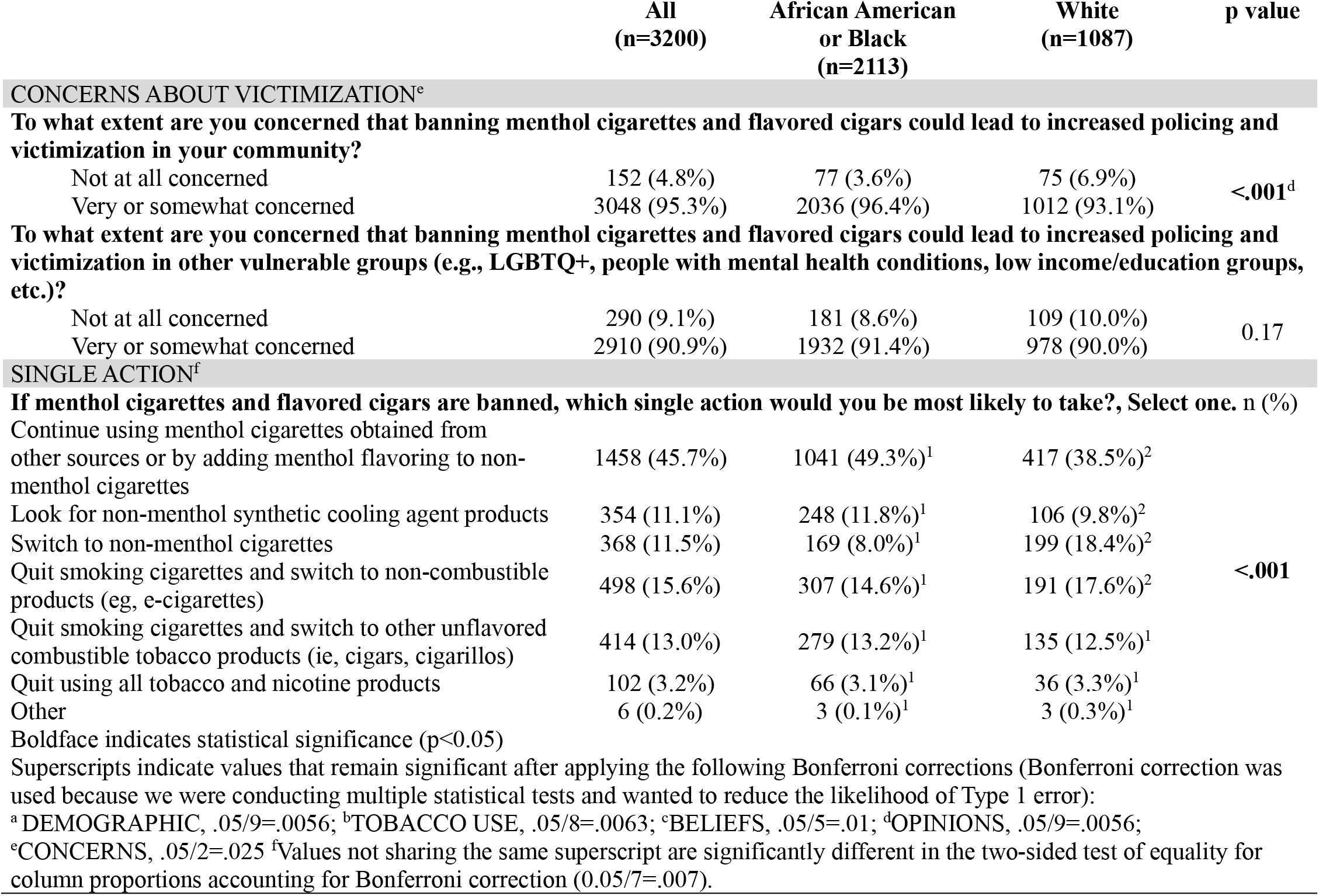
Comparison of Concerns about Victimization and Impact of a MC/FC Ban on Tobacco Use Behaviors by Race among AWS Menthol Cigarettes.

#### Concerns about victimization

Participants rated their concern that a MC/FC ban would lead to increased policing and victimization in their community and in communities of vulnerable groups, including Black people, LGTBQ+ people, people with mental health conditions and those with low income or education.

#### Impact on smoking behavior

Participants were asked what single action they would take if MC and FC were no longer available. Options included ‘continue using menthol cigarettes obtained from other sources or by adding menthol flavoring to non-menthol cigarettes,’ ‘look for non-menthol synthetic cooling agent products,’ ‘switch to non-menthol cigarettes,’ ‘quit smoking cigarettes and switch to non-combustible products (e.g., e-cigarettes),’ ‘quit smoking cigarettes and switch to other combustible tobacco products,’ and ‘quit using all tobacco and nicotine products.’ ^27^

### Statistical Analysis

Continuous variables were summarized with means and standard deviations. Categorical variables were summarized with frequencies and percentages. Independent samples t-tests with Bonferroni correction for continuous variables and Chi-square for categorical variables compared differences by race and ban favor versus opposition/ambivalence. Only contrasts that remained statistically significant after Bonferroni correction were considered statistically significant. To identify factors associated with ban opposition/ambivalence, variables that were significant between individuals who favored the ban versus those who opposed or were ambivalent to the ban at p<0.10 were entered into a stepwise logistic regression model. Only variables significant at p<0.05 were retained in the final model predicting ban opposition/ambivalence. A data-driven approach was used to ensure the best fit of models to the data. Sensitivity analyses used backward elimination logistic regression modeling to confirm the final model. Both stepwise and backward elimination procedures led to a final model comprised of the same eight factors. Results from the stepwise logistic regression model are presented.

Modeling opposition/ambivalence to a MC/FC ban allowed us to identify all participants not “in favor” who are an important target for public health messaging to increase public support of a federal product standard. All analyses were completed using SAS version 9.4 (SAS Institute Inc).

## RESULTS

Demographic and tobacco use characteristics of the sample are shown in Table 1.

### Race differences in perceptions and perceived impact of MC/FC ban

Black participants were more likely than Whites to have heard about a federal MC/FC ban (91.5% vs 79.7%, p<0.001); however, there was no statistical difference in favor versus opposition/ambivalence to a MC/FC ban with 37.2% of Black and 34.5% of White AWS menthol cigarettes indicating favor for a ban (p=0.13). Beliefs that menthol cigarettes are more addictive, harmful, and harder to quit were low overall [mean 1.3 (SD=1.1) out of 3.0 statements endorsed], but Black participants were more likely than White participants to endorse these beliefs [mean 1.4 statements (SD=1.1) versus 1.0 (SD=1.0), p<.001]. A greater proportion of Black adults also supported FDA’s rationale of the public health benefits of a MC/FC ban [3.0 statements (SD=1.7) versus 2.4 (SD=1.8) out of 5 statements endorsed, p<.001]. Both groups were concerned that a MC/FC ban would lead to increased policing and victimization in their community (95.3% overall) and other vulnerable communities (90.9% overall), although Black participants were more likely to endorse these as concerns in their community (96.4% vs 93.1% in White menthol users, p<0.001).

Both groups selected ‘continue using menthol cigarettes…’ as the most likely single action they would take if MC/FC were banned (45.7% overall); however, post-hoc analyses identified significant differences by race. Black AWS were more likely to look for ways to continue purchasing menthol cigarettes or by adding menthol flavoring to non-menthol cigarettes (B: 49.3%, W: 38.5%) and to look for non-menthol synthetic cooling agent products (B: 11.8%, W: 9.8%), while White AWS were more likely to switch to non-menthol cigarettes (W: 18.4% v B: 8.0%) or non-combustible products (W: 17.6% v B: 14.6%) (p<.007 all contrasts). Very few participants in either group endorsed quitting all tobacco and nicotine products as their most likely single action (3.2% overall).

### Ban Opposition/Ambivalence

Ban opposition/ambivalence did not differ statistically by race (Table 2). Primary reasons for being in favor (n=1,161) or opposed (n=1,122) are displayed in Supplementary Table 1. Those indicating ambivalence (n=917) were not queried about why they were ambivalent.

Univariate differences in favor versus opposition/ambivalence of a MC/FC ban across all variables are displayed in Supplementary Table 2. Twenty-three variables were significant at p<0.10, and eight factors remained in the final regression modeling ban opposition/ambivalence (Table 4). Owning a home (OR=0.59, 95% CI, 0.43-0.82), using OTP (OR=0.59, 95% CI, 0.38-0.91), using MJ (OR=0.52, 95% CI, 0.40-0.68), familiarity with a ban (OR=0.65, 95% CI, 0.49-0.86) and believing that their community would favor a ban (OR=0.37, 95% CI, 0.34-0.44) *decreased* the odds of opposition/ambivalence to a ban, while smoking more CPD (OR=1.04, 95% CI, 1.02-1.06) and belief that menthol cigarettes were more addictive, harmful, and harder to quit than non-menthol (OR=1.2, 95% CI, 1.01-1.3) *increased* the odds of ban opposition/ambivalence and intent to continue using menthol cigarettes. Further, those who selected a single action other than quitting all nicotine and tobacco products (i.e., selected ‘continue smoking menthol cigarettes obtained from other sources’) had *increased* odds of being opposed to a ban (see Table 4 for associated OR and 95% CI for each single action).

**Table 4.** Final stepwise logistic regression modeling factors associated with opposition/ambivalence to a MC/FC product standard.

| **Factors** | **OR (95% CI)** | **P Value** |
| --- | --- | --- |
| Home ownership (REF=No) |  |  |
| Yes | 0.59 (0.43-0.81) | 0.0012 |
| Cigarettes per day in the last 7 days | 1.04 (1.02-1.06) | <.0001 |
| OTP<sup>a</sup> use in the last 7 days (REF=No) |  |  |
| Yes | 0.59 (0.38-0.91) | 0.016 |
| MJ<sup>b</sup> use in the last 7 days (REF=No) |  |  |
| Yes | 0.52 (0.40-0.68) | <.0001 |
| Menthol cigarettes more addictive, harmful, and difficult to quit <sup>c</sup> | 1.18 (1.09-1.27) | <.0001 |
| Familiarity with product standard (REF= No) |  |  |
| Yes | 0.65 (0.49-0.86) | 0.0024 |
| Community perception of product standard (REF=opposed/ambivalent) |  |  |
| In favor | 0.37 (0.31-0.44) | <.0001 |
| Single action taken if a MC/FC product standard is enacted (REF=Quit using all tobacco and nicotine products entirely) |  | .004 |
| Continue menthol cigarettes obtained from other sources or by adding menthol flavoring to non-menthol cigarettes | 1.84 (1.17-2.89) |  |
| Look for non-menthol synthetic cooling agent products | 1.20 (0.74-1.96) |  |
| Quit smoking cigarettes and switch to combustible OTP | 1.56 (0.97-2.53) |  |
| Quit smoking cigarettes and switch to a non-combustible product (ie, e-cigarette or nicotine pouch) | 1.55 (0.97-2.50) |  |
| Switch to non-menthol cigarettes | 1.85 (1.13-3.03) |  |

Abbreviations: OTP, other tobacco product; MJ, marijuana; REF, reference; OR, odds ratio.
<sup>a</sup> Other tobacco product use was measured by e-cigarette, nicotine pouch, cigarillo, cigar, pipe, dip, and/or hookah utilization in the past 7 days.
<sup>b</sup> Marijuana use was measured by blunt, joint, spliff, bowl, bong, edible, and/or vaped marijuana use in the past 7 days.
<sup>c</sup> Continuous variable measuring participant endorsement of menthol cigarettes as more addictive, harmful, and difficult to quit (range 0-3) than non-menthol cigarettes.

## DISCUSSION

The current study compared perceptions of a menthol cigarette and flavored cigar (MC/FC) ban among Black and White adults who smoke (AWS) menthol cigarettes and examined factors associated with opposition or ambivalence to a ban. A primary finding is that support for a MC/FC ban did not differ statistically by race, with the proportion in favor close to 40%. This proportion is higher than the 28.5% support found among menthol smokers (race/ethnicity not specified) in a national survey conducted in 2021^16^ and indicate that public support for government action on this issue has grown in recent years. Support among Black participants in this study contradict recent headlines that cite fear of backlash from the Black community as one reason for failure to enact a MC/FC product standard^13^ and support actions by leaders of the Black community, the NAACP, and health organizations to promote the advancement of a MC/FC ban.^10,14-15^

Notably, Black participants demonstrated significantly greater support for the public health benefits of a ban than White participants. Specifically, Black participants were more likely to cite public health benefits, namely preventing youth from ever starting smoking, encouraging menthol smokers to smoke less, encouraging menthol smokers to quit, fewer smoking-related deaths, and improving the health of communities most impacted by menthol smoking as ‘likely’ outcomes of a MC/FC ban. Black participants were also more likely to endorse menthol cigarettes as addictive, harmful, and difficult to quit, suggesting that public health messaging targeting communities where menthol cigarette use is most widespread is being heard.^2-3,9,17^

Findings also indicate several important areas for future action. Specifically, most participants (63.7% total) remained opposed or ambivalent to a MC/FC product standard, indicating opportunity to increase support for a MC/FC ban among AWS menthol cigarettes. In this study, AWS menthol cigarettes who smoked more CPD, believed that menthol cigarettes were more addictive, difficult, and harder to quit, and did not intend to quit all nicotine and tobacco products had increased odds of opposition/ambivalence to a MC/FC ban. Findings suggest that messages targeting more addicted/dependent AWS menthol cigarettes could increase overall support for a MC/FC ban. Further, support for a menthol ban is more likely when respondents have accurate perceptions of the health consequences and addictive properties of menthol cigarettes and flavored cigars.^23^ Conversely, participants who believed that their community would be in favor of a MC/FC product standard and who were familiar with the ban reported *decreased* opposition. Black participants (91.5%) were more likely than White participants (79.7%, p<.001) to have heard about the ban, indicating that ongoing messaging to populations disproportionately impacted by menthol has the potential to decrease overall opposition. Findings from Sterling (2024) indicate widespread misperceptions about tobacco industry engagement around menthol policies and targeted communication that corrects this disinformation could decrease opposition to a MC ban.^17,30^ Overwhelming concern in our study about increased policing and victimization (95.3%, overall) of MC/FC users presents a clear opportunity to better clarify the intended outcomes from implementation of these policies.^31^

One of the intended outcomes of a MC/FC ban is to encourage menthol smokers to quit,^8,10-11,21,31-32^ yet only 3.2% of participants in our sample selected ‘quit using all tobacco and nicotine products’ as their likely course of action. This 3.2% is much lower than the 25-64% of US smokers who indicated that they would hypothetically attempt to quit smoking under a proposed ban in earlier studies^9,32^ but consistent with actual rates of 3.6% in a 2024 study among US menthol smokers ages 18-34 living under a real-world ban.^19^ The low proportion of participants indicating intent to quit in the face of a MC/FC ban is concerning, although actual response to an implemented ban could be higher.^33^ White respondents in the current study were more likely to indicate intent to switch to non-combustible tobacco products, consistent with literature that electronic nicotine delivery systems (ENDS) product users are predominantly White.^34^ Interest in non-menthol synthetic cooling agent products (i.e., WS-3, WS-23) as a substitute for menthol cigarettes was low (11.1%), but higher among Blacks (11.8%) than Whites (9.8%) and is a trend worth monitoring as the tobacco industry seeks regulatory loopholes to a MC/FC product standard.^35^

Limitations include use of MTURK as a crowdsourcing tool for participant recruitment. By virtue of using internet-based recruitment, older adults who use menthol cigarettes are underrepresented in the study population due to the skewed demographic of MTURK participants toward younger ages.^36^ Other MTURK limitations include convenience sampling, which does not reflect the demographics of the US population but is consistent with other studies in this area,^17-19,21,30,35^ reliance on self-report that is subject to reporting bias, and concerns for data quality control, which we mitigated using validity questions interspersed throughout the survey. Findings cannot be generalized to individuals from other racial/ethnic groups. Finally, participants were asked their opinions about the menthol cigarette and flavored cigar bans together rather than as separate policies.

## CONCLUSIONS

Over one-third of AWS menthol cigarettes were in favor of a MC/FC product standard, with no difference in support found between Black and White participants. Black participants demonstrated significantly greater support for the public health benefits of a MC/FC ban and were more likely than White participants to endorse menthol cigarettes as addictive, harmful, and difficult to quit. Targeted messaging to menthol users who smoke more CPD, believe that menthol cigarettes are more addictive, harmful, and harder to quit, and those who do not intend to quit all nicotine and tobacco products in response to a MC/FC product standard may increase support for a MC/FC ban. Overwhelming concern about increased policing/victimization of MC/FC users represents a clear opportunity to correct misperceptions about ban implementation and garner support. While a MC/FC product standard was withdrawn from the Office of Management and Budget register on January 21, 2025, momentum for implementing MC/FC bans continue to increase at the state and local level.^26^ In 2023 and 2024, 10 US states and over 400 municipalities introduced bills to ban the sale of menthol cigarettes. This is in addition to the 2 states and over 170 US cities with bans in place, suggesting relevance of the study findings irrespective of a federal ban on menthol cigarettes.^26^

## Data Availability

Data available: Yes
Additional Information: We will make the data and associated documentation available to users only under a data-sharing agreement that provides for: (1) a commitment to using the data only for research purposes and not to identify any individual participant; (2) a commitment to securing the data using appropriate computer technology; and (3) a commitment to destroying or returning the data after analyses are completed. All requests for data will go through a data request committee which will include Drs. Nollen and Mayo who will review any requests and will oversee data sharing procedures. If the request is deemed reasonable by the committee, we will proceed with a data sharing agreement and secure data sharing plan, using a secure file transfer system which meets industry and government regulations (e.g., HIPPA, GLBA, SOX) to ensure data security and compliance.
How to access data:
When available: With publication

## ACKNOWLEDGMENTS

We are grateful for the time and effort of Tricia Snow and her assistance with data collection and MTURK navigation.

## AUTHOR CONTRIBUTIONS

**Arnold:** Conceptualization, Methodology, Validation, Investigation, Resources, Data Curation, Writing – Original Draft, Writing – Review & Editing, Visualization, Supervision, Project Administration, Funding Acquisition. **Leavens:** Conceptualization, Methodology, Validation, Investigation, Writing – Review & Editing, Visualization, Project Administration. **Sanderson Cox:** Validation, Writing – Review & Editing. **Brown:** Methodology, Software, Validation, Formal Analysis, Resources, Data Curation, Writing – Review & Editing. **Mayo:** Conceptualization, Methodology, Software, Validation, Formal Analysis, Resources, Data Curation, Writing – Review & Editing. **Baldwin:** Writing – Original Draft, Writing – Review & Editing, Project Administration. **Nguyen:** Investigation, Resources, Data Curation. **Nollen:** Conceptualization, Methodology, Validation, Investigation, Resources, Data Curation, Writing – Original Draft, Writing – Review & Editing, Visualization, Supervision, Project Administration.

## STATEMENTS AND DECLARATIONS

### Ethics Approval Statement

The Institutional Review Board at the University of Kansas Medical Center approved our surveys (approval: STUDY00160222) on July 10, 2023. Respondents gave written consent for review and signature before starting interviews.

## DATA AVAILABILITY

### Data available

Yes

#### Additional Information

We will make the data and associated documentation available to users only under a data-sharing agreement that provides for: (1) a commitment to using the data only for research purposes and not to identify any individual participant; (2) a commitment to securing the data using appropriate computer technology; and (3) a commitment to destroying or returning the data after analyses are completed. All requests for data will go through a data request committee which will include Drs. Nollen and Mayo who will review any requests and will oversee data sharing procedures. If the request is deemed reasonable by the committee, we will proceed with a data sharing agreement and secure data sharing plan, using a secure file transfer system which meets industry and government regulations (e.g., HIPPA, GLBA, SOX) to ensure data security and compliance.

#### When available

With publication

