## Supplemental Figure 1 for "Perceptions of a Menthol Cigarette and Flavored Cigar Ban in Black and White Adults in the United States who Smoke Menthol Cigarettes and Factors Associated with Ban Opposition or Ambivalence"

159 with incomplete survey data

1,677 excluded based on inclusion criteria

- Live in a place or municipality where menthol is already banned (n=835)
- Non-menthol smoker (n=485)
- Primary user of other form(s) of tobacco (n=406)
- Not currently smoking cigarettes (n=224)
- Did not identify as Black or White (n=183)
- <21 years of age (n=176)
- Smoked menthol cigarettes for <1 year (n=20)

99 failed validity questions

9 with age outside feasible range (101-883)

2,133 identify as Black

1,087 identify as White

3,200 final analytic sample

3,308 with complete survey data

3,467 eligible after screening

5,144 workers completed screening

6,924 workers screened for eligibility

1,790 started but did not complete screening form
