## Supplemental Table 1 for "Perceptions of a Menthol Cigarette and Flavored Cigar Ban in Black and White Adults in the United States who Smoke Menthol Cigarettes and Factors Associated with Ban Opposition or Ambivalence"

| Supplementary Table 1. Participant’s primary reason for favor or opposition to the proposed FDA MC/FC product standard^a^ | | |
| --- | --- | --- |
|  | Frequency (n) | Percent (%) |
| IN FAVOR (n=1,161) | | |
| A ban will encourage me to quit smoking completely | 244 | 21.0 |
| A ban will reduce the number of smoking-related deaths  and improve public health | 226 | 19.5 |
| A ban will help the communities most impacted by  menthol smoking, especially Black people, LGBT+  people, people with mental health conditions, and those  with lower income or education | 226 | 19.5 |
| A ban will encourage me to reduce the amount that I  smoke | 209 | 18.0 |
| A ban will prevent youth from starting smoking | 126 | 10.9 |
| Menthol cigarettes are more harmful and/or addictive than  non-menthol cigarettes | 82 | 7.1 |
| Tobacco companies have historically targeted Black  communities with menthol cigarettes; a ban on menthol  cigarettes promotes health equity and social justice for the  Black community | 46 | 4.0 |
| Other | 2 | 0.2 |
| OPPOSED (n=1,122) | | |
| It is not the government’s place to tell me what I can and  cannot consume | 363 | 32.4 |
| It is just another example of a policy that unfairly targets  certain communities, especially Black people, LGBT+  people, people with mental health conditions, and those  with lower income or education | 296 | 26.4 |
| A ban would leave me no or few options for what to  smoke | 208 | 18.5 |
| The ban will not work- it could harm menthol smokers  because they will seek out other channels to buy menthol  cigarettes | 144 | 12.8 |
| There is no evidence that menthol cigarettes are more  harmful or addictive than menthol cigarettes | 71 | 6.3 |
| There is no evidence that menthol cigarettes encourage  vulnerable populations to smoke | 35 | 3.1 |
| Other | 5 | 0.4 |
| ^a^All study participants (n=3,200) were asked to indicate their favor, opposition, or ambivalence to the proposed MC/FC product standards. Only those who indicated favor (n=1,161) or opposition (n=1,122) were queried about their primary reason. The 917 who indicated ambivalence to a MC/FC product standard were not queried about their primary reason for being ambivalent. | | |
