## Supplemental Table 2 for "Perceptions of a Menthol Cigarette and Flavored Cigar Ban in Black and White Adults in the United States who Smoke Menthol Cigarettes and Factors Associated with Ban Opposition or Ambivalence"

| Supplementary Table 2. Factors associated with favor or opposition/ambivalence to a product standard prohibiting menthol cigarettes and flavored cigars | | | | |
| --- | --- | --- | --- | --- |
|  | **All  (n=3200)** | **Opposed or Ambivalent to Product Standard**  **(n=2039)** | **In Favor of Product Standard**  **(n=1161)** | **p value** |
| DEMOGRAPHIC CHARACTERISTICS^a^ | | | | |
| **Age,** mean (SD) | 33.9 (9.4) | 34.4 (9.8) | 33.0 (8.6) | <.001^a^ |
| **Age, years** n (%) |  |  |  | <.001 ^a^ |
| <40 | 2610 (81.6%) | 1626 (79.7%) | 984 (84.8%) |  |
| ≥ 40 | 590 (18.4%) | 413 (20.3%) | 177 (15.3%) |  |
| **Gender Identity,** n (%) |  |  |  | .88 |
| Female | 857 (26.8%) | 552 (27.1%) | 305 (26.3%) |  |
| Male | 2335 (73.0%) | 1482 (72.7%) | 853 (73.5%) |  |
| Transgender | 8 (0.3%) | 5 (0.3%) | 3 (0.3%) |  |
| **Race, n (%)** |  |  |  | .13 |
| African American or Black | 2113 (66.0%) | 1327 (65.1%) | 786 (67.7%) |  |
| White | 1087 (34.0%) | 712 (34.9%) | 375 (32.3%) |  |
| **Sexual Orientation,** n (%) |  |  |  | .027 |
| Straight, that is, not gay | 2505 (78.3%) | 1621 (79.5%) | 884 (76.1%) |  |
| Lesbian, Gay, Bisexual, Other | 695 (21.7%) | 418 (20.5%) | 277 (23.9%) |  |
| **Marital Status,** n (%) |  |  |  | <.001 ^a^ |
| Married or member of an unmarried couple | 2796 (87.4%) | 1747 (85.7%) | 1049 (90.4%) |  |
| Single, Divorced/Separated, Widowed | 404 (12.6%) | 292 (14.3%) | 112 (9.7%) |  |
| **Employment Status,** n (%) |  |  |  | .026 |
| Employed full or part-time | 3096 (96.8%) | 1962 (96.2%) | 1134 (97.7%) |  |
| Unemployed, Retired, Other | 104 (3.3%) | 77 (3.8%) | 27 (2.3%) |  |
| **Education Level,** n (%) |  |  |  | .078 |
| HS graduate, HS equivalent (GED), or less | 623 (19.5%) | 378 (18.5%) | 245 (21.1%) |  |
| Some college or more | 2577 (80.5%) | 1661 (81.5%) | 916 (78.9%) |  |
| **Income,** median (range) | $50,000  ($0-$155,000,000) | $50,000 ($0-$15,500,000) | $50,000 ($1-$155,000,000) | .77 |
| **Housing,** n (%) |  |  |  | <.001 ^a^ |
| Own a home | 2884 (90.1%) | 1782 (87.4%) | 1102 (94.9%) |  |
| Do not own a home | 316 (9.9%) | 257 (12.6%) | 59 (5.1%) |  |
| Tobacco Use Characteristics^b^ | | | | |
| **Age when you started smoking regularly,** mean (SD) | 15.7 (7.7) | 16.0 (7.6) | 15.0 (7.8) | <.001^b^ |
| **Length of time as a smoker in years,** mean (SD) | 18.2 (11.1) | 18.3 (11.3) | 18.0 (10.8) | .40 |
| **Length of time as a menthol smoker in years,** mean (SD) | 9.0 (7.4) | 9.2 (7.8) | 8.5 (6.8) | .003^b^ |
| **Cigarettes per day in the past 7 days,** mean (SD) | 7.9 (6.8) | 8.6 (7.8) | 6.8 (4.4) | <.001^b^ |
| **Nicotine Dependence,** **“How soon after waking do you first smoke,”** n (%) |  |  |  | <.001^b^ |
| Within 5 minutes | 577 (18.0%) | 421 (20.6%) | 156 (13.4%) |  |
| After 5 minutes | 2623 (82.0%) | 1618 (79.4%) | 1005 (86.6%) |  |
| **Other tobacco product use in the past 7 days,** n (% yes) | 2963 (92.6%) | 1834 (90.0%) | 1129 (97.2%) | <.001^b^ |
| Menthol flavored OTP (among OTP users) | 1885 (63.6%) | 1258 (68.6%) | 627 (55.5%) | <.001^b^ |
| **Marijuana use in the past 7 days,** n (% yes) | 2674 (83.6%) | 1609 (78.9%) | 1065 (91.7%) | <.001^b^ |
| BELIEFS ABOUT MENTHOL CIGARETTES AND PRIMARY REASON FOR USE^c^ | | | | |
| **Addictiveness of menthol vs. non-menthol cigarettes,** n (%) |  |  |  | <.001^c^ |
| Less or equally addictive | 1658 (51.8%) | 1000 (49.1%) | 658 (56.7%) |  |
| More addictive | 1541 (48.2%) | 1038 (50.9%) | 503 (43.3%) |  |
| **Harmfulness of menthol vs. non-**  **menthol cigarettes,** n (%) |  |  |  | <.001^c^ |
| Less or equally as harmful | 1941 (60.7%) | 1186 (58.2%) | 755 (65.0%) |  |
| More harmful | 1258 (39.3%) | 852 (41.8%) | 406 (35.0%) |  |
| **Difficulty to quit of menthol vs. non-menthol cigarettes,** n (%) |  |  |  | <.001^c^ |
| Less or equally as hard to quit | 1928 (60.3%) | 1145 (56.2%) | 783 (67.4%) |  |
| Harder to quit | 1271 (39.7%) | 893 (43.8%) | 378 (32.6%) |  |
| **Overall beliefs about Menthol Summary: Out of the 3 statements regarding beliefs about menthol cigarettes, how many thought menthol was more addictive, more harmful, and harder to quit,** mean (SD) | 1.3 (1.1) | 1.4 (1.1) | 1.1 (1.0) | <.001^c^ |
| **What is the primary reason you smoke menthol cigarettes?,** n (%) |  |  |  | <.001^c^ |
| They taste better | 1088 (34.0%) | 805 (39.5%) | 283 (24.4%) |  |
| They are less harsh, smoother, or easier to smoke | 831 (26.0%) | 534 (26.2%) | 297 (25.6%) |  |
| They are less harmful than non-menthol cigarettes | 623 (19.5%) | 347 (17.0%) | 276 (23.8%) |  |
| They are what my family and friends smoke | 387 (12.1%) | 196 (9.6%) | 191 (16.5%) |  |
| They are heavily advertised to me and my  community | 163 (5.1%) | 84 (4.1%) | 79 (6.8%) |  |
| They are easier to get and more available to me  than non-menthol cigarettes | 95 (3.0%) | 63 (3.1%) | 32 (2.8%) |  |
| They are cheaper than non-menthol cigarettes | 12 (0.4%) | 9 (0.4%) | 3 (0.3%) |  |
| Other | 1 (0.0%) | 1 (0.1%) | 0 (0.0%) |  |
| OPINIONS OF THE PROPOSED MENTHOL AND FLAVORED CIGAR PRODUCT STANDARD^d^ | | | | |
| **Before today, had you heard about the FDA’s proposed ban on menthol cigarettes and all flavored cigars?,** n (%) |  |  |  | <.001^d^ |
| Yes | 2799 (87.5%) | 1729 (84.8%) | 1070 (92.2%) |  |
| No | 401 (12.5%) | 310 (15.2%) | 91 (7.8%) |  |
| **In your opinion, how would others in your community feel about the proposed ban on menthol cigarettes and flavored cigars?,** n (%) |  |  |  | <.001^d^ |
| Opposed or ambivalent | 1883 (58.8%) | 1416 (69.5%) | 467 (40.2%) |  |
| In favor | 1317 (41.2%) | 623 (30.6%) | 694 (59.8%) |  |
| **How likely do you think each of the following things is to happen if the FDA bans menthol cigarettes and flavored cigars?,** n (%) |  |  |  |  |
| **Prevent youth from ever starting smoking** |  |  |  | .61 |
| Unlikely outcome of the FDA ban | 1101 (34.4%) | 695 (34.1%) | 406 (35.0%) |  |
| Likely outcome of the FDA ban | 2099 (65.6%) | 1344 (65.9%) | 755 (65.0%) |  |
| **Encourage menthol smokers to smoke less** |  |  |  | .08 |
| Unlikely outcome of the FDA ban | 1336 (41.8%) | 828 (40.6%) | 508 (43.8%) |  |
| Likely outcome of the FDA ban | 1864 (58.3%) | 1211 (59.4%) | 653 (56.2%) |  |
| **Encourage menthol smokers to quit** |  |  |  | .44 |
| Unlikely outcome of the FDA ban | 1487 (46.5%) | 937 (46.0%) | 550 (47.4%) |  |
| Likely outcome of the FDA ban | 1713 (53.5%) | 1102 (54.1%) | 611 (52.6%) |  |
| **Lead to fewer smoking-related deaths and improve public health** |  |  |  | .20 |
| Unlikely outcome of the FDA ban | 1500 (46.9%) | 938 (46.0%) | 562 (48.4%) |  |
| Likely outcome of the FDA ban | 1700 (53.1%) | 1101 (54.0%) | 599 (51.6%) |  |
| **Improve the health of communities most impacted by menthol smoking, including Black, LGBTQ+, people with mental health conditions, low income/education groups, etc.)** |  |  |  | .79 |
| Unlikely outcome of the FDA ban | 1531 (47.8%) | 972 (47.7%) | 559 (48.2%) |  |
| Likely outcome of the FDA ban | 1669 (52.2%) | 1067 (52.3%) | 602 (51.9%) |  |
| **Overall support for FDA rationale: Out of the 5 statements regarding potential outcomes of a ban on menthol cigarettes and flavored cigars, how many were endorsed as very or somewhat likely,** mean (SD) | 2.8 (1.8) | 2.9 (1.8) | 2.8 (1.7) | .20 |
| CONCERNS about victimization^e^ | | | | |
| **To what extent are you concerned that banning menthol cigarettes and flavored cigars could lead to increased policing and victimization in your community?** |  |  |  | .01^e^ |
| Not at all concerned | 152 (4.8%) | 111 (5.4%) | 41 (3.5%) |  |
| Very or somewhat concerned | 3048 (95.3%) | 1928 (94.6%) | 1120 (96.5%) |  |
| **To what extent are you concerned that banning menthol cigarettes and flavored cigars could lead to increased policing and victimization in other vulnerable groups (eg, LGBTQ+, people with mental health conditions, low income/education groups, etc.)?** |  |  |  | .78 |
| Not at all concerned | 290 (9.1%) | 187 (9.2%) | 103 (8.9%) |  |
| Very or somewhat concerned | 2910 (90.9%) | 1852 (90.8%) | 1058 (91.1%) |  |
| WHAT SINGLE ACTION WOULD YOU TAKE^f^ | | | | |
| **If menthol cigarettes and flavored cigars are banned, which single action would you be most likely to take?** n (%) |  |  |  | <.001 |
| Continue using menthol cigarettes obtained from other sources or by adding menthol flavoring to non-menthol cigarettes | 1458 (45.7%) | 1013 (49.8%)^1^ | 445 (38.4%)^2^ |  |
| Look for non-menthol synthetic cooling agent products | 354 (11.1%) | 179 (8.8%) ^1^ | 175 (15.1%) ^2^ |  |
| Switch to non-menthol cigarettes | 368 (11.5%) | 260 (12.8%) ^1^ | 108 (9.3%) ^2^ |  |
| Quit smoking cigarettes and switch to non-combustible products (e.g., e-cigarettes) | 498 (15.6%) | 295 (14.5%) ^1^ | 203 (17.5%) ^2^ |  |
| Quit smoking cigarettes and switch to other combustible tobacco products | 414 (13.0%) | 233 (11.5%) ^1^ | 181 (15.6%) ^2^ |  |
| Quit using all tobacco and nicotine products | 102 (3.2%) | 55 (2.7%) ^1^ | 47 (4.1%) ^2^ |  |
| Other | 6 (0.2%) | 4 (0.2%) ^1^ | 2 (0.2%) ^1^ |  |
| Superscripts indicate values that remain significant after applying the following Bonferroni corrections:  ^a^ DEMOGRAPHIC, .05/10=.005; ^b^TOBACCO USE, .05/8=.0063; ^c^BELIEFS, .05/5=.01; ^d^OPINIONS, .05/8=.0071; ^e^CONCERNS, .05/2=.025  ^f^Values not sharing the same superscript are significantly different in the two-sided test of equality for column proportions accounting for Bonferroni correction (0.05/7=.007). | | | | |
